# Quantification and prognostic significance of interferon-γ secreting SARS-CoV-2 responsive T cells in hospitalised patients with acute COVID-19

**DOI:** 10.1101/2021.09.21.21263902

**Authors:** Daniel Pan, Jee Whang Kim, Joshua Nazareth, Sara Assadi, Adam Bellass, Jack Leach, James G Brosnan, Adam Ahmed, Fleur Starcevic, Shirley Sze, Christopher A Martin, Caroline M Williams, Michael R Barer, Amandip Sahota, Prashanth Patel, Andrea Tattersall, Andrea Cooper, Manish Pareek, Pranabashis Haldar

**Affiliations:** Department of Respiratory Sciences, University of Leicester, United Kingdom; Department of Infectious Diseases and HIV Medicine, University Hospitals of Leicester NHS Trust, United Kingdom; Respiratory Biomedical Research Centre, University of Leicester, United Kingdom; University of Leicester Medical School, United Kingdom; Department of Cardiovascular Sciences, University of Leicester, United Kingdom; Department of Microbiology, University Hospitals of Leicester NHS Trust, United Kingdom; Department of Chemical and Metabolic Diseases, University Hospitals of Leicester NHS Trust, United Kingdom; Oxford Immunotec Ltd, United Kingdom; Trudeau Institute, Saranac Lake, New York, United States of America

**Author notes:** Joint first authorship (Daniel Pan, Jee Whang Kim, Joshua Nazareth). Joint senior authorship (Manish Pareek, Pranabashis Haldar).

## Abstract

Little is known about T-cell responses during acute coronavirus disease-2019 (COVID-19). We measured T-cell interferon gamma (IFN-γ) responses to spike 1 (S1), spike 2 (S2), nucleocapsid (N) and membrane (M) SARS-CoV-2 antigens using the T-SPOT® Discovery SARS-CoV-2 assay, a proven EliSPOT technology, in 114 hospitalised adult COVID-19 patients and assessed their association with clinical disease phenotype. T-SPOT® Discovery SARS-CoV-2 responses were detectable within 2 days of a positive PCR and did not correlate with vaccination status or symptom duration. Higher responses to S1 protein associated with a higher symptom burden, and serum IL-6 levels. Despite treatment with dexamethasone this subgroup was also at greater risk of requiring continuous positive airway pressure (CPAP) in the days following sampling. Higher T-cell responses measured using T-SPOT® Discovery SARS-CoV-2 associate with progressive disease in acute COVID-19 disease and may have utility as a prognostic biomarker that should be evaluated in larger cohorts.

## Introduction

The clinical assessment of patients presenting to hospital with Coronavirus Disease-2019 (COVID-19) can be difficult. In particular, patients can appear to be well on initial assessment, but deteriorate rapidly during their hospital stay.^1^ Measurement of the T-cell response during acute COVID-19 may help to predict disease severity.^2^ Current studies have demonstrated high frequency of SARS-COV-2-specific T-cell responses in individuals who have recovered from severe COVID-19 compared those who had mild COVID-19.^3^ T-cell immunity has also been found to be important in long-term protection against SARS-CoV-2 and studies indicate dissociation between T-cell and neutralizing antibody responses during convalescence. However, the T-cell response during acute disease and its association with humoral immunity remains poorly characterized.

Here we investigated the systemic T-cell response during acute SARS-CoV-2 infection in a hospitalised cohort of patients with COVID-19, using a functional T-cell assay developed to measure T-cell responses to four antigenic domains of SARS-CoV-2. We hypothesized that systemic T-cell responses in acute COVID-19 disease would be heterogeneous and associate with interval from symptom onset, prior vaccination status and disease severity.

## Results

### Patient recruitment and characteristics

Between 8^th^ February and 8^th^ March 2021, 114 patients were prospectively recruited, of which 104 patients were admitted to hospital with a primary diagnosis of acute COVID-19 disease. The remaining 10 patients acquired SARS-CoV-2 in hospital. These participants were negative on nasopharyngeal PCR swab at the time of hospital admission for other reasons, and developed a positive PCR during their hospital stay that was identified as part of screening undertaken for infection control. Table 1 shows participant demographic data. The median age was 64 (IQR 52-78). Most participants were male (n=69, 61%) and of White ethnicity (n=89, 79%). 102 (91%) were symptomatic (fever, cough, breathlessness, anosmia) at time of sampling. 88 (79%) had infiltrates on their admission chest x-ray. 31% of patients had received one dose of either the Pfizer BioNTech or Oxford AstraZeneca vaccine (n=36, 31%) in the weeks prior to acute infection; 29 had received their vaccine 2 weeks or longer prior to admission. In keeping with the UK vaccine policy, this was an older aged cohort. The median duration of symptoms prior to sampling was 10 days (IQR 7 to 15). Most patients had been treated with dexamethasone (n=73, 64%); Almost all patients were antibody positive at time of sampling (n=95, 93%).

### Association between T-SPOT panels and antibody responses

Valid T-SPOT assay readings were obtained in 87 patients (76%). In the remainder, the assay could not be performed due to an insufficient number of peripheral blood mononuclear cells in the samples collected. These patients were more lymphopenic (median lymphocyte count in 0.8×10^9^ cells/ml (IQR 0.4 to 1.1); compared with 1.3 ×10^9^ cells/ml (IQR 1.0 to 1.7), p<0.001). In the 87 subjects with a measurable T-cell response, the responses to the spike protein antigens S1 and S2 were most sensitive, being positive in the highest proportion of participants (Table 3) and at the greatest amplitude. The median T-SPOT for S1 was 5 spots (IQR 2 to 54); S2: 5 spots (IQR 2 to 22); Nucleocapsid: 3 spots (IQR 0 to 7); Membrane: 3 spots (IQR 1 to 10). Table 2 shows the associations between different T-cell panels and combined IgG/IgM for the S1. Strong correlation was observed between the response to S1 and responses to the other three antigens. However, there was little concordance between T-SPOT responses and the antibody assay. We observed no association of T-cell responses with either prior vaccination status or interval after symptoms onset. The T-SPOT assay was positive in the 12 asymptomatic patients and could be detected within 3 days of a positive PCR test. In the 10 nosocomial patients, T-SPOT assays were also detectable 16 days after an initially positive PCR test.

### Relationship between T-SPOT responses and markers of disease severity

84 (73%) participants received oxygen during hospitalisation; a fifth required continuous positive airway pressure (CPAP) in the days following blood sampling (n=24, 21%) for progressive respiratory failure. None required mechanical ventilation. 7 (6%) study participants died within 28 days of hospital admission. Table 3 shows the relationship between clinical variables and responses to all four antigens of the T-SPOT assay. In general, patients with higher T-cell responses to S1 protein were more likely to be symptomatic, have raised IL-6 levels, receive dexamethasone for COVID-19 and receive CPAP prospectively (Figure 1).

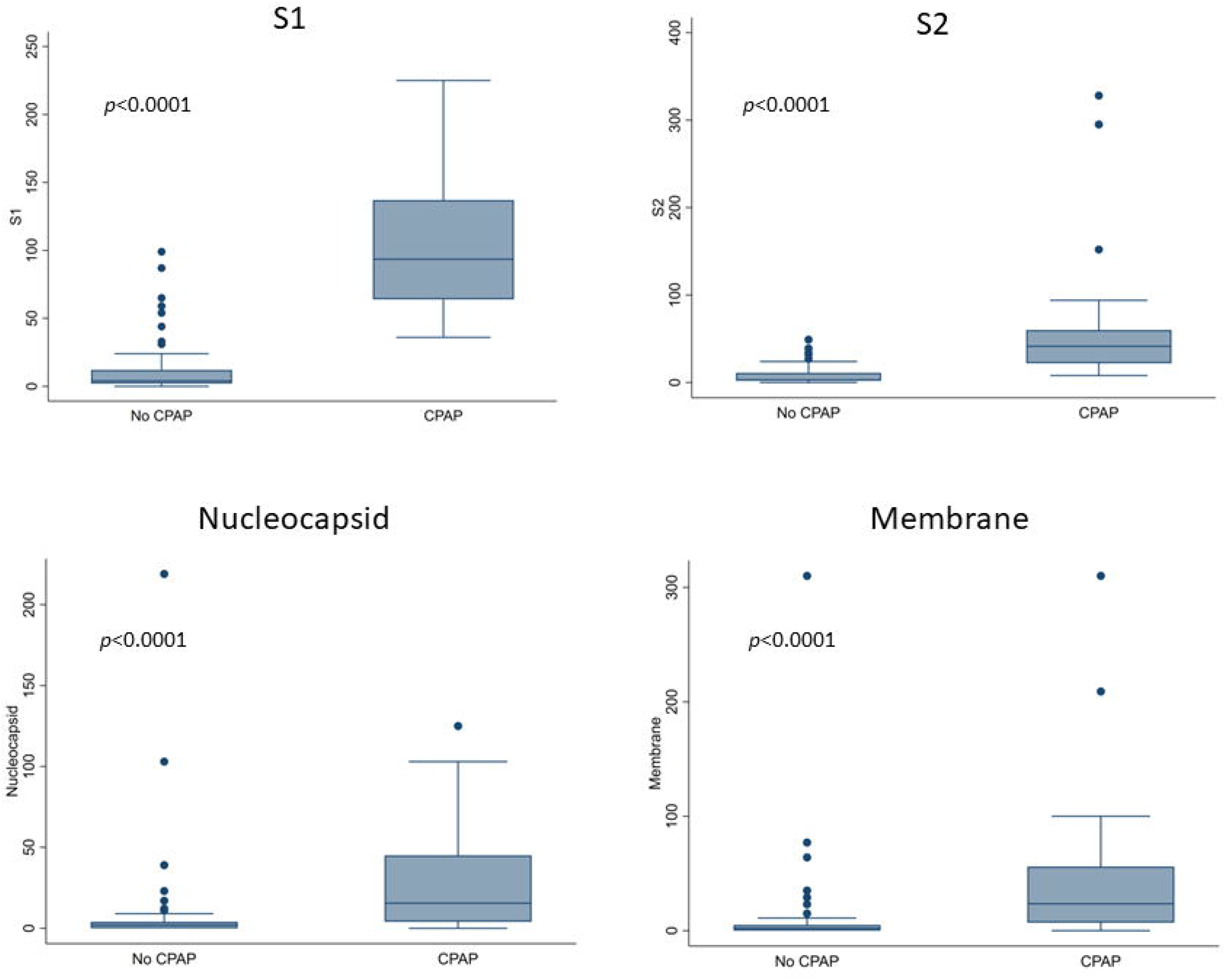

## Discussion

Our study is the first to evaluate T-cell responses using the T-SPOT assay in hospitalised patients with acute COVID-19. We found that T-cell responses appeared as early as two days after symptom onset in early COVID-19, and can be positive in the context of a negative combined antibody assay. Second, in a hospitalised cohort, we found T-cell responses to be similar between vaccinated and unvaccinated participants. Finally, we found that higher T-cell responses, predominantly those to S1 appear to be related to a more severe disease phenotype.

Previous studies have assessed the utility of the T-SPOT assay in convalescent patients and found that T-SPOT responses to the same proteins in this study were seen in the absence of anti-Spike IgG on long-term follow-up. We found similar levels of discordance in patients with acute COVID-19, highlighting the potential of the T-SPOT assay to pick up immunological responses in COVID-19 positive patients where antibody responses are negative. T-SPOT responses were also much higher in studies in convalescent individuals, highlighting clear differences in the kinetics of the T-cell response compared with antibody response over time after infection and vaccination.^4,5^

We provide real-world data on the T-cell response of patients who had received one dose of the Pfizer BioNTech or Oxford AstraZeneca vaccine prior to developing COVID-19 requiring hospitalisation. We did not observe amplification of the T-cell response in this group, compared with the unvaccinated population. This could reflect the design of current vaccines, which focus on generating a neutralizing antibody response rather than T-cell response, measurement of T-cell responses in a primarily older cohort where there is immunosenescence or may reflect insufficient time for the vaccines to induce robust T-cell immunity, despite the fact that the majority of our patients had their first dose 2 weeks or longer prior to admission

The presence of early T-cell responses during acute COVID-19 has been shown in smaller cohorts using different assays. Tan and colleagues examined the dynamic changes in the immunological response in 12 patients with COVID-19 and also found both antibody responses and interferon-γ-SARS-CoV-2 specific T cells to be present as early as 2 days from symptom onset^6^, in keeping with the findings in this study. It appears that immune activation after infection with SARS-CoV-2 occurs rapidly, before the virus can evade or suppress recognition, even in the absence of previous infection or vaccination.

We found an association between higher T-SPOT responses (especially S1) and increasing disease severity at the time of sampling, as evidenced by higher IL-6 levels and prospective need for CPAP. It is not clear whether these T-cell responses are protective or deleterious. A protective role for the higher T-cell responses in severe disease is supported by the low 28-day mortality rate in this cohort. However, it is possible that the elevated T-cell responses are a marker of immune hyperstimulation generating cytokine over-production and cell death.^7^ Studies comparing T cell phenotype and cytokine levels are needed to resolve this question.

Our study was limited by size and sociodemographic heterogeneity, with a variable proportion exposed to immunomodulatory interventions, including dexamethasone treatment and a prior history of receiving one dose of a two-dose vaccination regimen. In contrast to studies on convalescent patients, a quarter of samples from our cohort could not be processed to perform the T-SPOT assay due to inadequate cell numbers and is likely a consequence of the lymphopenia that occurs in acute COVID-19; future studies using the assay can overcome this by taking larger quantities of blood. Despite this, our study demonstrates that T-cell responses to the S1 antigen, measured using the T-SPOT assay appear early in infection and associate with objective measures of disease severity - these observations warrant further investigation in other cohorts. Our findings also support the need for larger studies to investigate the kinetics of T-cell responses in acute infection and to build understanding of how these responses influence short and long term prognosis.

## Methods

### Participant recruitment

We conducted a prospective observational cohort study of hospitalised and nosocomially infected adult patients at University Hospitals of Leicester NHS Trust. Patients were eligible if they were 16 years or over, tested positive for SARS-COV-2 on nasopharyngeal RT-PCR using the hospital assay, with no previous history or record of infection and no existing conditions or treatments associated with T cell immunodeficiency.

### Blood sampling

One 10ml lithium heparin anticoagulated blood for T-cell functional assay and one 6ml blood anticoagulated using EDTA to measure antibody response was taken from each study participant within 24 hours of a positive routine PCR test. Serology was performed using the commercially available SARS-CoV-2 Total Assay manufactured by Siemens, which detects IgG and IgM to the S1 RBD antigen and gave a qualitative result.^8^

### T-SPOT assay

To measure T cell responses, we used the T-SPOT® Discovery SARS-CoV-2 kit (T-SPOT), which uses an ELISpot technology to detect IFN-γ release from T cells after exposure to four SARS-CoV-2 peptides antigens: Spike protein S1 and S2 domains, Membrane and Nucleoprotein peptides. ^9^ In brief, cells were isolated and washed from peripheral blood and SARS-CoV-2 responsive T cell numbers are enumerated using a T-SPOT Discovery SARS-CoV-2 tests, which are based on the same proven analytical platform as the T-SPOT®.TB (tuberculosis) kit also manufactured by Oxford Immunotec. Peripheral blood mononuclear cells (PBMCs) were treated with T-Cell Xtend® reagent (Oxford Immunotec Ltd) prior to isolated from whole blood.

T cells isolated from whole blood stored overnight appear to show reduced responses to stimulation with antigens in ELISPOT assays, but this is primarily due to contaminating cell populations in the PBMC layer. The T-Cell Xtend reagent contains bispecific monoclonal antibodies which are directed against cell surface markers on selected white blood cells and glycophorin A on the red blood cells. The T-Cell Xtend reagent cross-links the selected white blood cells with the red blood cells, which increases the density of the selected cells. When a density gradient is applied during FICOLL extraction, the selected white blood cells remain separated in the red blood cell layer away from the PBMC layer. Non-selected cells, including T cells and antigen presenting cells, are contained in the PBMC layer.

Briefly, 10 mL of blood was treated with 250 μL T-Cell Xtend reagent After addition of reagent, tubes were inverted five times and left to stand at room temperature for 20 minutes before starting the isolation of PBMCs. PBMCs were isolated by standard ficoll-hypaque density-gradient centrifugation from heparinized blood samples. The PBMC were washed twice with RPMI-1640 medium, counted with an automated haematology analyser and adjusted in AIM V medium such that each of four wells of the assay plate contained 250,000 PBMC.

The cells were incubated at 37 °C and 5% CO2 for 16–20 h with medium (negative control), phytohemagglutinin (positive control) and peptides. After 16-20 hours incubation, the wells were washed and developed using a conjugated secondary antibody that bound to any IFN-γ captured on the membrane. After washing to remove unbound IFN-γ, substrate was added to produce dark spots of insoluble precipitate indicating areas of IFN-γ secretion from T cells. These spot forming cells (SFCs) were counted using an automated ELISpot plate reader (CTL, Shaker Heights, OH) and manually verified.

Each subject had to display a satisfactory response to the positive control (> 20 spots and/or saturation of the well) and a low spot number in the negative control (≤ 10 spots of the well); otherwise, the test was classified as indeterminate.

### Clinical data and ethical approval

Routine clinical, radiological, laboratory and demographic data at the time of sampling was collected and prospective outcomes during admission including requirement for CPAP, invasive ventilation and 28 day mortality, were recorded. The study had ethical approval from the West Midlands Research Ethics Committee (REC Reference 20/WM/0153).

### Statistical analysis

Continuous variables are expressed as median and interquartile range (IQR). Categorical variables are displayed as numbers and percentages (%). Pearson’s Chi-squared and Fisher’s exact row test was used to compare categorical variables between groups. Student’s t-test and Kruskal-Wallis test were used to compare continuous variables between groups depending on the normality of distribution. We chose to examine the relationship between increasing T-SPOT responses in the four panels separately in relation to clinical variables. The association between different T-SPOT panels was assessed using Pearson’s correlation coefficient; association between T-SPOT panels and antibody response was assessed using the Kappa statistic.

All analyses were performed using STATA version 14.2 (StataCorp United States) and Excel version 2016 (Microsoft, Redmond, United States). A p-value <0.05 was considered to be statistically significant.

## Data Availability

Data is available upon reasonable request.

## Notes

### Competing Interest Statement

The authors have declared no competing interest.

### Clinical Trial

REC Reference 20/WM/0153

### Funding Statement

Not applicable - no external funding was received for this work.

### Author Declarations

The study had ethical approval from the West Midlands Research Ethics Committee (REC Reference 20/WM/0153).

## References

1. Sze, S. et al. Variability, but not admission or trends in NEWS2 score predicts clinical outcome in elderly hospitalised patients with COVID-19. J. Infect. 82, 159–198 (2021).

2. Sabbaghi, A. et al. Role of γδ T cells in controlling viral infections with a focus on influenza virus: implications for designing novel therapeutic approaches. Virol. J. 17, 1–18 (2020).

3. Sekine, T. et al. Robust T Cell Immunity in Convalescent Individuals with Asymptomatic or Mild COVID-19. Cell 183, 158–168.e14 (2020).

4. Wyllie, D. et al. SARS-CoV-2 responsive T cell numbers and anti-Spike IgG levels are both associated with protection from COVID-19: A prospective cohort study in key workers. medrxi (2021).

5. Clarke, C. L. et al. Longevity of SARS-CoV-2 immune responses in hemodialysis patients and protection against reinfection. Kidney Int. 99, 1470–1477 (2021).

6. Tan, A. T. et al. Early induction of functional SARS-CoV-2-specific T cells associates with rapid viral clearance and mild disease in COVID-19 patients. Cell Rep. 34, (2021).

7. McElvaney, O. J., Curley, G. F., Rose-John, S. & McElvaney, N. G. Interleukin-6: obstacles to targeting a complex cytokine in critical illness. Lancet Respir. Med. 9, 643–654 (2021).

8. Ainsworth, M. et al. Performance characteristics of five immunoassays for SARS-CoV-2: a head-to-head benchmark comparison. Lancet Infect. Dis. 20, 1390–1400 (2020).

9. Abubakar, I. et al. Two interferon gamma release assays for predicting active tuberculosis: The UK PREDICT TB prognostic test study. Health Technol. Assess. (Rockv). 22, 1–95 (2018).

