## Supplementary material for "Quantification and prognostic significance of interferon-γ secreting SARS-CoV-2 responsive T cells in hospitalised patients with acute COVID-19": Tables 1, 2, 3a and 3b

| Table 1: Participant demographic, clinical, laboratory and radiological data and clinical outcomes | | |
| --- | --- | --- |
| **Variables** | **Patients** | **Missing data** |
| **Demographic data** | | |
| Age – median years (IQR) | 64 (52 to 78) | 0 |
| Male – n (%) | 69 (61%) | 0 |
| White ethnicity – n (%)  Asian ethnicity – n (%)  Black ethnicity - n (%) | 89 (79%)  23 (20%)  2 (1%) | 0 |
| Autoimmune disease – n (%)  Hypertension – n (%)  Diabetes – n (%)  Ischaemic heart disease – n (%)  Chronic kidney disease – n (%)  Cancer – n (%)  Chronic lung disease – n (%)  Neurological disease – n (%)  Gastroenterological/liver disease – n (%)  Haematological- n (%)  Number of comorbidities – median (IQR) | 20 (18%)  42 (37%)  30 (26%)  35 (31%)  9 (8%)  5 (4%)  23 (20%)  13 (11%)  10 (9%)  5 (4%)  1 (1-2) | 0 |
| **Clinical data** | | |
| Admission oxygen saturations – median % (IQR)  Any oxygen in hospital – n (%) | 96% (94 to 97)  84 (73%) | 0 |
| White cell count – median x10^9^ cells/L (IQR)  Lymphocyte - median x10^9^ cells/L (IQR)  Urea – median mmol/L (IQR)  Creatinine – median μmol/L (IQR)  CRP – median mg/L (IQR)  Haemoglobin – median g/L (IQR)  IL-6 – median pg/ml | 8.0 (6.2-11.0)  1.27 (0.87-1.68)  7.1 (5.1-9.6)  68 (57-89)  21 (5-60)  129 (113-141)  23 (10-68) | 13 (not performed on sampling)  13  13  13  13  13  86 (never performed) |
| Nosocomial acquired infection – n(%) | 10 (8%) | 0 |
| Findings of COVID-19 pneumonia on CXR – n (%) | 88 (79%) | 2 (CXR never performed) |
| Duration of symptoms – median days (IQR) | 10 (7 to 15) | 0 |
| Treatment with dexamethasone – n (%) | 73 (64%) | 0 |
| Vaccinated - n (%)  Pfizer – n (%)  Astra Zeneca – n (%) | 36 (31%)  24 (21%)  12 (11%) | 0 |
| Combined IgG/IgM positive– n (%) | 95 (93%) | 12 inconclusive |
| T cell responses – median spots (IQR)  Panel 1/S1 protein  Panel 2/S2 protein  Panel 3/Nucleocapsid protein  Panel 4/Membrane protein | 5 (2 to 54)  5 (2 to 22)  3 (0 to 7)  3 (1 to 10) | 27 inconclusive |
| **Clinical outcomes** |  |  |
| Received CPAP following sampling – n (%) | 24 (21%) | 0 |
| 28 day mortality – n (%) | 7 (6%) | 0 |

| Table 2: Association between T-SPOT panels and combined S-protein IgG/IgM against SARS-CoV-2. Pearson’s correlation coefficient (and *p* values) is displayed for association between T-SPOT panels, and Kappa statistic (and *p* values) are displayed for association between T-spot panels and antibody assay. | | | | |
| --- | --- | --- | --- | --- |
|  | **S2** | **Nucelocapsid** | **Membrane** | **Antibody** |
| **S1** | 0.54  (<0.001) | 0.32  (0.003) | 0.51  (<0.001) | 0.06  (0.27) |
| **S2** |  | 0.19  (0.08) | 0.39  (<0.001) | 0.09  (0.18) |
| **Nucleocapsid** |  |  | 0.50  (<0.001) | 0.17  (0.01) |
| **Membrane** |  |  |  | 0.11  (0.08) |

| Table 3a: Clinical variables by increasing T cell response to S1 and S2 protein. Categorical variables are expressed as number (%); continuous variables are expressed as median (interquartile range, IQR). | | | | | |
| --- | --- | --- | --- | --- | --- |
| **Variables** | **0 (n=12)** | **1-5 (n=32)** | **6-54 (n=22)** | **>54 (n=21)** | ***P* value** |
| **S1 protein** | | | | | |
| Age - years | 66 (55 to 79) | 62(47 to 76) | 65 (56 to 75) | 75 (56 to 81) | 0.51 |
| Male | 5 (42%) | 23 (72%) | 9 (41%) | 13 (62%) | 0.08 |
| White ethnicity | 10 (83%) | 25 (78%) | 18 (82%) | 17 (81%) | 0.99 |
| Number of comorbidities | 0 (0 to 1) | 1 (0 to1) | 1 (0 to 1) | 1 (0 to 1) | 0.84 |
| Any oxygen in hospital | 1 (1 to2) | 2 (1 to 3) | 1 (1 to 2) | 2 (1 to 2) | 0.65 |
| Received NIV | 0 (0%) | 0 (0%) | 1 (5%) | 17 (81%) | <0.001 |
| White cell count - x10^9^ cells/L  Lymphocyte - x10^9^ cells/L  Urea - mmol/L  Creatinine - μmol/L  CRP - mg/L  Haemoglobin - –g/L  IL-6 - pg/ml | 6.7 (5.1 to 12.2)  1.2 (0.9 to 1.5)  7.3 (2.6 to 9.0)  63 (55 to 70)  5 (5 to 26)  134 (98 to 144)  12 (6 to 23) | 9.5 (6.7 to 11.2)  1.2 (1.1 to 1.6)  6.5 (5.0 to 9.3)  67 (60 to 96)  26 (9 to 84)  128 (118 to 142)  18 (5 to 38) | 8.3 (6.2 to 11.0)  1.3 (0.9 to 1.6)  5.7 (5.1 to 8.8)  65 (56 to 86)  16 (5 to 65)  128 (115 to 133)  13 (9 to 85) | 7.2 (6.2 to 9.7)  1.6 (1.3 to 2.4)  8.2 (6.0 to 10.2)  72 (60 to 84)  11 (7 to 26)  119 (104 to 146)  450 (100 to 558) | 0.66  0.09  0.28  0.38  0.23  0.78  0.04 |
| Findings of COVID pneumonia on CXR | 9 (75%) | 23 (74%) | 20 (91%) | 16 (76%) | 0.44 |
| Vaccinated | 6 (50%) | 11 (34%) | 7 (28%) | 6 (29%) | 0.37 |
| Antibody negative | 1 (8%) | 2 (7%) | 0 (0%) | 1 (5%) | 0.68 |
| Symptomatic | 9 (75%) | 26 (81%) | 22 (100%) | 20 (95%) | 0.04 |
| Days since symptom onset at time of sampling | 9 (7-12) | 11 (7-15) | 10 (8-17) | 12 (9-17) | 0.44 |
| Dexamethasone | 6 (50%) | 17 (53%) | 19 (86%) | 14 (67%) | 0.04 |
| Received CPAP following sampling | 0 (0%) | 0 (0%) | 1 (5%) | 17 (81%) | <0.001 |
| 28 day mortality | 0 (0%) | 1 (3%) | 2 (9%) | 1 (5%) | 0.82 |
| **S2 protein** | **0 (n=10)** | **1-7 (n=36)** | **8-27 (n=24)** | **>27 (n-17)** | ***P* value** |
| Age – years | 66 (54 to 74) | 64 (49 to 80) | 65 (55 to 82) | 72 (55 to 78) | 0.82 |
| Male | 6 (60%) | 22 (61%) | 12 (50%) | 10 (59%) | 0.85 |
| White ethnicity | 8 (80%) | 29 (81%) | 20 (83%) | 13 (77%) | 0.96 |
| Number of comorbidities | 0 (0 to 1) | 1 (0 to1) | 1 (0 to 1) | 1 (0 to 1) | 0.84 |
| Any oxygen in hospital | 1 (1 to 2) | 2 (1 to 3) | 1 (1 to 3) | 1 (1 to 2) | 0.67 |
| White cell count - x10^9^ cells/L  Lymphocyte - x10^9^ cells/L  Urea - mmol/L  Creatinine - μmol/L  CRP - mg/L  Haemoglobin - –g/L  IL-6 - pg/ml | 8.9 (5.1 to 10.9)  1.4 (0.9 to 1.5)  7.6 (2.9 to 8.5)  63 (55 to 70)  5 (5 to 53)  134 (94 to 141)  12 (6 to 23) | 7.6 (6.2 to 11.3)  1.1 (0.9 to 1.4)  6.6 (4.6 to 9.7)  75 (56 to 98)  26 (9 to 84)  127 (115 to 139)  18 (5 to 27) | 8.5 (5.9 to 11.1)  1.4 (1.3 to 2.1)  6.4 (5.5 to 9.6)  65 (56 to 85)  14 (5 to 47)  130 (109 to 141)  29 (10 to 79) | 9.2 (6.8 to 11.4)  1.6 (1.3 to 2.4)  8.1 (5.1 to 10.0)  72 (68 to 82)  11 (7 to 23)  121 (113 to 142)  275 (100 to 450) | 0.96  0.02  0.46  0.18  0.43  0.99  0.15 |
| Findings of COVID pneumonia on CXR | 7 (70%) | 27 (77%) | 20 (83%) | 14 (82%) | 0.84 |
| Vaccinated | 4 (40%) | 13 (36%) | 6 (25%) | 6 (35%) | 0.78 |
| Antibody negative | 1 (11%) | 2 (6%) | 1 (4%) | 0 (0%) | 0.58 |
| Symptomatic | 7 (70%) | 31 (86%) | 23 (96%) | 16 (94%) | 0.17 |
| Days since symptom onset at time of sampling | 8 (6 to 9) | 12 (7 to 17) | 10 (8 to 17) | 11 (8 to 13) | 0.18 |
| Dexamethasone | 5 (50%) | 20 (56%) | 19 (79%) | 12 (71%) | 0.20 |
| Received CPAP following sampling | 0 (0%) | 0 (0%) | 5 (21%) | 13 (76%) | <0.001 |
| 28 day mortality | 0 (0%) | 2 (6%) | 1 (4%) | 1 (6%) | 0.99 |

| Table 3b: Clinical variables by increasing T cell response to Nucleocapsid and Membrane protein. Categorical variables are expressed as number (%); continuous variables are expressed as median (interquartile range, IQR). | | | | | |
| --- | --- | --- | --- | --- | --- |
| **Variables** | **0 (n=22)** | **1-2 (n=18)** | **3-5 (n=22)** | **>5 (n=24)** | ***P* value** |
| **Nucleocapsid protein** | | | | | |
| Age - years | 66 (54 to 76) | 64 (52 to 83) | 63 (47 to 78) | 68 (57 to 76) | 0.97 |
| Male | 12 (55%) | 13 (72%) | 11 (48%) | 14 (58%) | 0.46 |
| White ethnicity | 19 (86%) | 16 (89%) | 17 (74%) | 18 (75%) | 0.55 |
| Number of comorbidities | 2 (1 to 2) | 1 (0 to 3) | 1 (1 to 2) | 1 (1 to 2) | 0.73 |
| Any oxygen in hospital | 14 (64%) | 14 (78%) | 18 (78%) | 15 (63%) | 0.53 |
| White cell count x10^9^ cells/L  Lymphocyte x10^9^ cells/L  Urea –mmol/L  Creatinine –μmol/L  CRP –mg/L  Haemoglobin –g/L (IQR)  IL-6 –pg/ml | 8.1 (6.5 to 10.9)  1.3 (1.1 to 1.5)  6.1 (4.6 to 8.8)  64 (52 to 70)  32 (5 to 84)  128 (101 to 140)  18 (7 to 36) | 8.6 (6.2 to 11.2)  1.4 (0.9 to 2.1)  7.0 (5.4 to 9.8)  83 (60 to 96)  11 (5 to 62)  134 (127 to 153)  68 (35 to 322) | 9.9 (6.0 to 13.4)  1.3 (1.0 to 1.6)  6.2 (5.1 to 8.5)  70 (62 to 88)  19 (7 to 36)  129 (121 to 138)  18 (5 to 104) | 7.5 (6.2 to 11.0)  1.4 (1.2 to 2.4)  8.7 (4.8 to 10.2)  71 (56 to 105)  11 (7 to 45)  115 (103 to 134)  13 (7 to 17) | 0.70  0.77  0.70  0.28  0.99  0.20  0.26 |
| Findings of COVID pneumonia on CXR | 17 (77%) | 14 (78%) | 21 (95%) | 16 (67%) | 0.10 |
| Vaccinated | 8 (36%) | 5 (28%) | 10 (33%) | 6 (25%) | 0.40 |
| Antibody negative | 3 (14%) | 0 (0%) | 0 (0%) | 1 (4%) | 0.13 |
| Symptomatic | 16 (73%) | 17 (94%) | 22 (96%) | 22 (92%) | 0.11 |
| Days since symptom onset at time of sampling | 9 (5 to 13) | 11 (8 to 19) | 11 (9 to 17) | 11 (7 to 15) | 0.26 |
| Dexamethasone | 11 (50%) | 14 (78%) | 18 (78%) | 13 (54%) | 0.10 |
| Received CPAP following sampling | 1 (5%) | 2 (11%) | 3 (13%) | 12 (50%) | 0.001 |
| 28 day mortality | 2 (9%) | 0 (0%) | 2 (9%) | 0 (0%) | 0.27 |
| **Membrane protein** | **0 (n=19)** | **1-2 (n=24)** | **3-10 (n=23)** | **>10 (n=21)** | ***P* value** |
| Age – years | 65 (54 to 76) | 69 (45 to 84) | 62 (45 to 78) | 64 (57 to 76) | 0.93 |
| Male | 12 (63%) | 15 (63%) | 12 (52%) | 11 (52%) | 0.81 |
| White ethnicity | 17 (89%) | 21 (88%) | 17 (74%) | 15 (71%) | 0.33 |
| Number of comorbidities | 1 (1 to 3) | 2 (1 to 2) | 1 (1 to 2) | 1 (1 to 2) | 0.92 |
| Any oxygen in hospital | 11 (58%) | 16 (67%) | 19 (83%) | 15 (71%) | 0.36 |
| White cell count x10^9^ cells/L  Lymphocyte x10^9^ cells/L  Urea –mmol/L  Creatinine –μmol/L  CRP –mg/L  Haemoglobin –g/L (IQR)  IL-6 –pg/ml | 8.4 (5.1 to 12.2)  1.3 (1.1 to 1.6)  6.5 (4.8 to 8.8)  67 (55 to 82)  12 (5 to 84)  131 (95 to 147)  17 (12 to 20) | 7.5 (5.9 to 11.2)  1.1 (0.9 to 1.6)  6.8 (4.4 to 9.7)  66 (60 to 93)  23 (5 to 74)  128 (105 to 139)  28 (25 to 55) | 9.9 (7.7 to 11.2)  1.3 (1.0 to 1.7)  8.2 (5.2 to 9.8)  72 (58 to 92)  20 (5 to 53)  130 (122 to 138)  44 (9 to 100) | 7.2 (6.8 to 10.4)  1.5 (1.3 to 2.5)  7.3 (5.3 to 10.1)  71 (60 to 93)  10 (7 to 25)  120 (111 to 139)  10 (3 to 49) | 0.52  0.22  0.64  0.77  0.75  0.98  0.54 |
| Findings of COVID pneumonia on CXR | 14 (78%) | 18 (75%) | 19 (83%) | 17 (81%) | 0.92 |
| Vaccinated | 7 (38%) | 9 (37%) | 7 (31%) | 6 (29%) | 0.61 |
| Antibody negative | 2 (11%) | 1 (4%) | 1 (5%) | 0 (0%) | 0.53 |
| Symptomatic | 15 (79%) | 19 (79%) | 23 (100%) | 20 (95%) | 0.03 |
| Days since symptom onset at time of sampling | 10 (6 to 14) | 9 (7 to 13) | 11 (8 to 17) | 12 (10 to 17) | 0.36 |
| Dexamethasone | 9 (47%) | 15 (63%) | 18 (78%) | 14 (67%) | 0.22 |
| Received CPAP following sampling | 1 (5%) | 3 (13%) | 2 (9%) | 12 (57%) | <0.001 |
| 28 day mortality | 2 (11%) | 1 (4%) | 1 (4%) | 0 (0%) | 0.49 |
